# A polygenic risk score improves the prediction of cardiovascular risk associated with obstructive sleep-apnea

**DOI:** 10.64898/2025.12.08.25341829

**Authors:** Christian W. Thorball, Adrien Waeber, Geoffroy Solelhac, Flavia Hodel, Théo Imler, Grégory Heiniger, Roxane de La Harpe, Pedro Marques-Vidal, Peter Vollenweider, Jacques Fellay, Raphael Heinzer

## Abstract

**Introduction:** Obstructive sleep apnea (OSA) carries increased cardiovascular (CV) risk. However, this risk is not fully captured by the apnea–hypopnea index (AHI). We investigated whether a validated coronary artery disease polygenic risk score (CAD-PRS) refines CV risk assessment in OSA.

**Methods:** We derived CAD-PRS using genome-wide genotyping data for 1,379 participants of the CoLaus|HypnoLaus cohort who underwent polysomnography. Associations between OSA, CAD-PRS, clinical factors, and incident CV events were assessed using multivariable Cox proportional hazards models. Risk stratification improvement was assessed with reclassification analyses compared to clinical risk scores (SCORE2/SCORE2-OP).

**Results:** During 7.2 years of median follow-up, 100 participants experienced CV events. A significant interaction between OSA and CAD-PRS was observed (p=0.013). The effect of OSA on CV risk differed across PRS categories. In the intermediate genetic-risk group (CAD-PRS quintiles 2–4), OSA patients (AHI ≥15/h) had a markedly higher CV risk compared to non-OSA (HR[95% CI]: 2.68[1.54–4.66]), whereas OSA did not significantly increase CV risk in either the low or high PRS strata. The complete model with OSA, CAD-PRS and their interaction allowed a significant reclassification (Net Reclassification Index 0.171, p=0.014) compared to SCORE2/SCORE2-OP and 52% of individuals at intermediate risk were reclassified as low or high CV risk.

**Conclusion:** A CAD-PRS refines CV risk stratification in individuals with OSA. The impact of OSA on CV risk is greatest in individuals with intermediate genetic risk. Adding CAD-PRS and OSA to SCORE2 improves model performance and reclassification, supporting more precise CV risk assessment in OSA.

**Statement of significance:** Obstructive sleep apnea is common, however the apnea–hypopnea index alone does not adequately capture associated cardiovascular risk. In this study, we evaluated whether a polygenic risk score for coronary artery disease can refine cardiovascular risk assessment in people with sleep apnea. Both sleep apnea and genetic risk were independently associated with incident cardiovascular events beyond standard clinical risk scores. Importantly, we found a significant interaction: excess cardiovascular risk related to moderate-to-severe sleep apnea was mainly confined to individuals with an intermediate genetic risk, while those at low or very high genetic risk were less affected. These findings support the potential value of incorporating genetic information to enable more nuanced and personalized cardiovascular risk assessment in sleep apnea.

## INTRODUCTION

Obstructive sleep apnea (OSA) is characterised by repetitive episodes of partial or complete airway obstruction during sleep and affects up to 49% of men and 23% of women over 40 years old^1^. OSA has been associated with various adverse health outcomes, including daytime sleepiness, increased risk of hypertension, stroke, heart failure, diabetes and metabolic syndrome^2–5^. Gold standard sleep recordings allow to accurately measure each of these respiratory events. Traditionally, OSA severity is defined according to the apnea-hypopnea index (AHI) by counting the number of respiratory events per hour of sleep^6^. Standard and rather arbitrary thresholds are used as an indication for treatment. However, randomised controlled trials suggest that the AHI may not be specific enough to single out patients with OSA at risk for developing apnea-associated cardiovascular diseases (CVD) and who would benefit from treatment^7–9^. This raises the need for more specific markers for cardiovascular (CV) risk stratification in OSA^10^.

In the last decade, several studies have tried to target patients at risk of apnea-associated CVD using specific polysomnographic biomarkers^11^ such as hypoxic burden^12^, respiratory events lengths^13^, sleep apnea-related pulse rate response^14,15^, pulse wave amplitude drop index^16^ or subjective complaints such as excessive daytime sleepiness^17^ or insomnia complaints^18,19^. However, even if sleep apnea is an independent cardiovascular risk factor, it remains essential to consider the patient’s overall cardiovascular risk profile, using validated global CV risk assessment tools, such as SCORE2/SCORE2-OP^20,21^ which have been developed to predict 10 years risk of first-onset CVD in European populations. In addition, integrating polygenic risk scores (PRS) and other genetic information holds promise for further enhancing cardiovascular risk stratification among patients with OSA, by capturing genetic susceptibility beyond traditional clinical factors^22^.

In that view, genome-wide association studies (GWAS) have enabled the development of PRS, which combine the effects of many genetic variants throughout the genome to effectively quantify the individual genetic predisposition to CVD^23^. Applying PRS to high-risk groups such as OSA patients is still debated. For example, in the UK Biobank, the effect of OSA on CAD risk did not change depending on a person’s CAD-PRS, but significant interactions emerged when using scores focused on specific pathways of intermittent hypoxemia^24^. This suggests that the genes and OSA interplay is unlikely to be uniform. Instead, it may involve specific biological pathways, meaning that a true interaction could be obscured when using broad, undifferentiated PRS models.

Therefore, the primary aim of the present study was to assess whether a genome-wide CAD-PRS enhances cardiovascular risk prediction in OSA and to specifically investigate the potential for a non-linear interaction between genetic predisposition and OSA severity. The secondary aim was to investigate whether integrating PRS within a validated clinical CV risk score (SCORE2/SCORE2-OP) improves risk reclassification in OSA patients. Identifying high-risk OSA patients using genetic testing may enable more personalized management and intensifying control of other cardiovascular risk factors.

## METHODS

### Ethics statement

The institutional Ethics Committee of the University of Lausanne, which afterwards became the Ethics Commission of Canton Vaud (www.cer-vd.ch) approved the CoLaus|PsyCoLaus study (project number PB_2018-00038, reference 239/09). All participants gave their signed informed consent before entering the study.

### Participants

CoLaus**|**PsyCoLaus is a prospective cohort study aiming to evaluate the prevalence and associations of mental disorders and cardiovascular risk factors in the community of Lausanne, Switzerland and to identify genetic determinants and mechanisms involved in their association. The sample, composed of 6733 people, was randomly selected from the 35 to 75-year-old residents of the city of Lausanne, Switzerland, from 2003 to 2006 according to the civil register and 4791 subjects participated in the genetic analysis^25^.

All participants underwent thorough physical and psychiatric evaluations at baseline and at three follow-up visits (every 5 years approximately). During first follow-up, 2162 participants (aged 45-80 years) took part in the sleep sub-cohort HypnoLaus, and underwent an unattended overnight full polysomnography (PSG) at home between 2009 and 2013^1^. Prospective data on cardiovascular events were obtained from the third follow-up of the CoLaus|PsyCoLaus study (between 2018 and 2021).

Inclusion criteria were: participation in the HypnoLaus study with valid PSG data, availability of genotyping data, no history of CVD at the time of PSG, availability of follow-up data, and European ancestry. As illustrated in Figure 1, a total of 1,379 individuals met these criteria and were included in the final analysis. Participants of non-European ancestry were excluded to ensure the validity of the CAD-PRS, which was developed and validated in populations of European descent.

**Figure 1.**
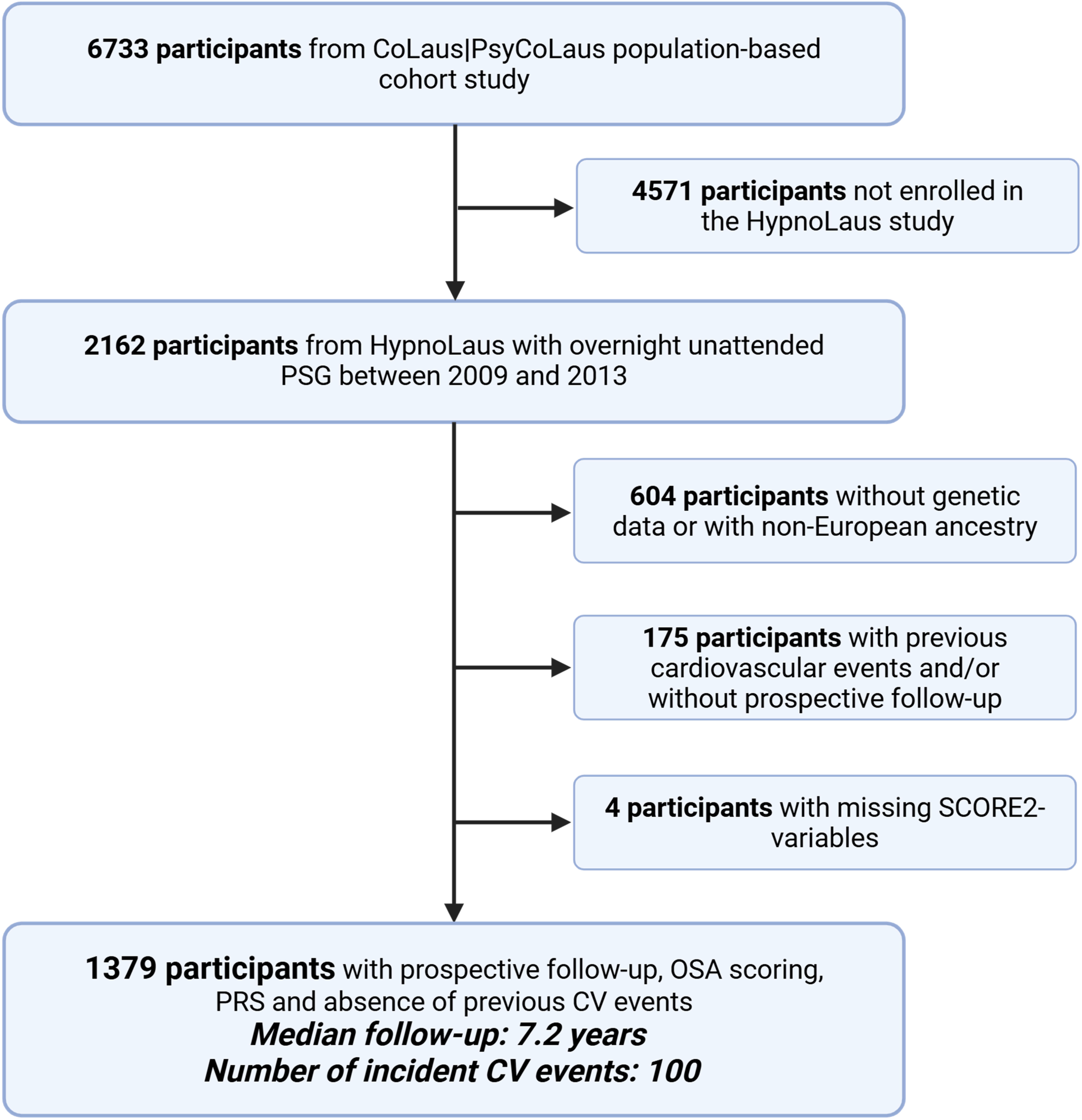
Study flow chart. Flow chart showing selection of participants from the CoLaus|PsyCoLaus cohort and the HypnoLaus sleep sub-study to the final analytic sample with polysomnography, CAD-PRS and follow-up. Participants with missing PSG or genetic data, prior cardiovascular events, non-European ancestry or no follow-up were excluded. The median follow-up was 7.2 years, during which 100 incident CV events occurred. Abbreviations: PSG, polysomnography; OSA, obstructive sleep apnea; CAD-PRS, coronary artery disease polygenic risk score; CV, cardiovascular

### Cardiovascular outcomes

The **primary outcome** was incident cardiovascular (CV) events, defined as a composite of cardiovascular death, stroke (definite or probable), acute myocardial infarction (AMI, definite or probable), and coronary heart disease (CHD, definite or probable).

For sensitivity analyses, we considered three alternative definitions of cardiovascular outcome:

1. **Hard CV outcomes (including transient ischemic attack (TIA)):** cardiovascular death, stroke (definite or probable) and TIA (excluding amaurosis fugax and transient global amnesia), AMI (definite or probable), and CHD (definite or probable).
2. **Hard CV outcomes (excluding TIA):** cardiovascular death, stroke (definite or probable; excluding TIA, amaurosis fugax and transient global amnesia), AMI (definite or probable), and CHD (definite or probable).
3. **Pure hard CV outcomes:** cardiovascular death, definite stroke, definite AMI, and definite CHD.

Events were identified through medical records, hospital databases, death registries, and follow-up interviews, and were independently adjudicated by at least two specialists (cardiologists, neurologists, or internists) using predefined international criteria, as described by Beuret et al.^26^.

Time-to-event was calculated from the date of PSG to the first incident CV event or the last follow-up, whichever occurred first.

### Clinical assessment

Blood samples were collected following overnight fasting for biochemical assays, including glucose, insulin and lipid profiles as previously described^25^. Smoking status, alcohol consumption, and use of medications were reported and sleep questionnaires were collected (Epworth Sleepiness Scale (ESS), Pittsburgh Sleep Quality Index (PSQI)).^27,28^

Anthropometric measurements were performed by trained observers with standard techniques. Body weight, height, and waist circumference were measured with participants standing without shoes in light clothes. Body mass index (BMI) was calculated as body mass in kg divided by the square of the patient’s height in meters.

BP was measured in triplicate on the left arm and values averaged between the last two readings. Arterial hypertension was defined as a systolic BP (SBP) ≥ 140 mmHg and/or a diastolic BP (DBP) ≥ 90 mmHg or, current use of antihypertensive medication.

### Polysomnography

The participants performed a full night PSG at home (Titanium, Embla® Flaga, Reykjavik, Iceland). PSG were performed according to the American Academy of Sleep Medicine (AASM) 2007 recommendations and included: EEG leads (F3, F4, C1, C2, O1 and O2, 256 Hz sampling rate), electrooculography (EOG, left and right), electromyography (EMG, chin and anterior tibialis muscle), electrocardiography (ECG, one lead), oxygen saturation (SpO2), airflow (nasal cannula), abdominal and thoracic respiratory efforts, snoring, and body position. PSG data were visually scored according to the AASM 2007 guidelines^29^.

Two trained sleep technicians scored polysomnographic recordings using Somnologica software (version 5.1.1, Embla Flaga, Reykjavik, Iceland). An expert sleep clinician reviewed every recording, and a second sleep expert did random quality checks. We defined apnea as a drop of at least 90% of airflow from baseline lasting 10 s or longer. We scored hypopnea events with 2012 AASM criteria^30^ (≥30% drop of airflow lasting at least 10 s with either an arousal or ≥3% oxygen saturation drop). We reported the average number of apnea and hypopnea events per hour of sleep (AHI). OSA status was defined as a binary variable (non-moderate/severe: AHI <15 events/h vs. moderate/severe: AHI ≥15 events/h).

### Genotyping and imputation

DNA samples were genotyped using the BB2 GSK-customized Affymetrix Axiom Biobank array. Variants and samples with excessive missingness above 5% or variants deviating from Hardy-Weinberg equilibrium (HWE, P < 10e-7) were excluded prior to imputation.

Imputation was performed in two separate runs with the Sanger Imputation Service^31^, using Positional Burrows Wheeler Transform (PBWT)^32^ with variants first phased using EAGLE2 (v2.0.5)^33^ and then imputed using the merged 1000 Genomes Project phase 3 plus UK10K reference panel and subsequently the Haplotype Reference Consortium r1.1 reference panel. The two imputed datasets were then merged, retaining only high-quality single nucleotide polymorphisms (SNPs) with an imputation information score (INFO) > 0.8. In cases where variants in both datasets had an INFO score above this threshold, the variant from the dataset with the highest INFO score was retained. For the final dataset, rare variants (minor allele frequency below 1%), variants with high missingness (above 10%) or with a deviation from HWE (P < 10e-7) were removed prior to calculating the PRS. Assessment of the presence of potential cryptic related or duplicate samples was performed using KING (v2.2.6)^34^. Ancestry was determined using principal component analysis with PLINK (v2.00a3LM)^35^ together with the 1000 Genomes Project reference panel.

### Polygenic risk score

The CAD-PRS was derived using PRSice (v2.3.5)^36^ based on the previously validated CAD-PRS by Inouye et al.^23^, with information on the included variants and their respective effect sizes obtained from the Polygenic Score Catalog^37^ (PGS000018). This PRS was developed using data from the UK Biobank, with CAD defined as diagnosis of fatal or non-fatal myocardial infarction (MI), or history of percutaneous transluminal coronary angioplasty (PTCA), or of coronary artery bypass grafting (CABG). In total, 1,357,679 of 1,745,179 variants from the original CAD-PRS were successfully matched and included in this study. The resulting raw PRS scores were standardized (i.e., scaled to a mean of 0 and a standard deviation of 1) prior to analysis.

### SCORE2/SCORE2-OP

SCORE2/SCORE2-OP risk scores were calculated at time of PSG to assess 10-year cardiovascular risk using the low-risk model as applicable to Switzerland using the RiskScorecvd (v0.3.1) R package^38^. Categories were defined by the published SCORE2/SCORE2-OP age-specific 10-year risk thresholds for fatal and non-fatal CVD (low, intermediate, high risk: <2.5%, 2.5–7.5%, ≥7.5% for individuals <50 years; <5%, 5–10%, ≥10% for 50–69 years; and <7.5%, 7.5–15%, ≥15% for ≥70 years)^20,21^.

### Statistical analyses

Baseline characteristics of the study population were summarized using medians and interquartile ranges (IQR) for continuous variables and frequencies with percentages for categorical variables. Differences in baseline characteristics between participants with and without incident CV events were assessed using the Wilcoxon rank-sum test for continuous variables and Pearson’s Chi-squared test for categorical variables.

The primary outcome was first incident CV event. The association between risk factors and incident CV events was evaluated using Cox proportional hazards models. The proportionality of hazards assumption was assessed graphically using log-log survival plots and formally tested using Schoenfeld residuals.

To evaluate the independent and combined predictive value of clinical risk, genetic risk, and OSA, a series of nested Cox models was constructed. The base model included the SCORE2/SCORE2-OP risk score as a continuous variable. The CAD-PRS was included as a categorical variable. We categorized CAD-PRS into three strata: Low (bottom quintile, <20%), Intermediate (quintiles 2–4, 20–80%), and High (top quintile, >80%). The Intermediate group served as the reference category. This stratification was chosen to maximize statistical power by establishing a large, stable reference group, and to specifically capture non-linear risk trajectories where clinical utility is concentrated in the tails of the genetic distribution, as previously described by Mega et al.^39^ and Marston et al.^40^ All models were adjusted for population stratification by including the top five principal components of the genotyping matrix as covariates.

To test for a statistical interaction between genetic risk and OSA, an interaction term (PRS group × OSA status) was added to the multivariable Cox model containing SCORE2, PRS, and OSA main effects. The significance of the interaction was determined using the likelihood ratio test (LRT).

Model performance was assessed using several metrics. The improvement in model fit for nested models was quantified using the LRT p-value, Akaike Information Criterion (AIC), and Bayesian Information Criterion (BIC). Discriminatory ability was evaluated using Harrell’s C-index. The proportion of variance explained was estimated using Nagelkerke’s pseudo-R2. Calibration was assessed numerically using the optimism-corrected calibration slope (200 bootstrap resamples) via the rms (v8.1-0) R package^41^. Visual calibration for the 10-year prediction horizon was evaluated using the riskRegression (v2025.09.17) R package^42^. Smoothed calibration curves were generated using nearest neighbour estimation with jackknife pseudo-values to account for censoring, superimposed with a density plot to illustrate the distribution of predicted risks.

To evaluate clinical utility and risk stratification improvement, reclassification analyses were performed. We calculated the Integrated Discrimination Improvement (IDI) and the continuous Net Reclassification Improvement (NRI) for models with the addition of PRS, OSA, and their interaction plus the five first principal components, compared to the base model using the survIDINRI R package^43^. The categorical NRI was determined using established age-dependent risk strata for SCORE2/SCORE-OP, and a bootstrap procedure with 1000 replicates was employed to derive its 95% confidence interval and p-value.

Adjusted cumulative incidence curves were generated from the final Cox model to visualize the probability of incident CV events over time across different strata of PRS and OSA status.

All statistical analyses were conducted using R (v4.4.2).

## RESULTS

After excluding 783 HypnoLaus participants due to missing genotyping data, non-European ancestry, prevalent cardiovascular disease at baseline, or other missing data points, 1,379 individuals were included in the analysis, of which 100 individuals experienced at least one adjudicated definite or probable CV event during follow-up (Figure 1). Baseline characteristics at the time of PSG are shown in Table 1.

Participants who suffered a CV event were more likely to have OSA, were older and more frequently diabetic. They also had higher BMI, higher systolic blood pressure and higher HDL cholesterol levels. They also exhibited more severe sleep-disordered breathing, characterized by higher AHI, higher oxygen desaturation index at 3% (ODI-3%), lower mean oxygen saturation, poorer sleep efficiency, and a lower pulse wave amplitude drop index (PWADi). Baseline characteristics of participants according to OSA status are shown in Table S1.

### CAD PRS is an independent risk factor for CVD among sleep apnea patients

We first evaluated whether the addition of the CAD-PRS to the validated SCORE2/SCORE2-OP algorithm improved the prediction of incident CV events among participants with and without moderate-to-severe OSA.

In Cox models adjusted for SCORE2/SCORE2-OP, both CAD-PRS and moderate-to-severe OSA were independently associated with CV risk (Figure 2). Using the lowest 20% of the CAD-PRS distribution as the reference, participants with PRS between the 20th and 80th percentile had a 1.57-fold higher risk of CV events (HR 1.57, 95% CI 0.86–2.89, p-value = 0.14), and those in the top 20% had an almost threefold higher risk (HR 2.88, 95% CI 1.50–5.53, p-value = 0.001). Similarly, moderate-to-severe OSA was associated with a 1.77-fold increased risk of CV events (HR 1.77, 95% CI 1.17–2.66, p-value = 0.007) compared with participants without OSA, independently of SCORE2. Each one-unit increase in SCORE2 was associated with a 14% higher risk of CV events (HR 1.14, 95% CI 1.11–1.18, p-value < 0.001).

**Figure 2.**
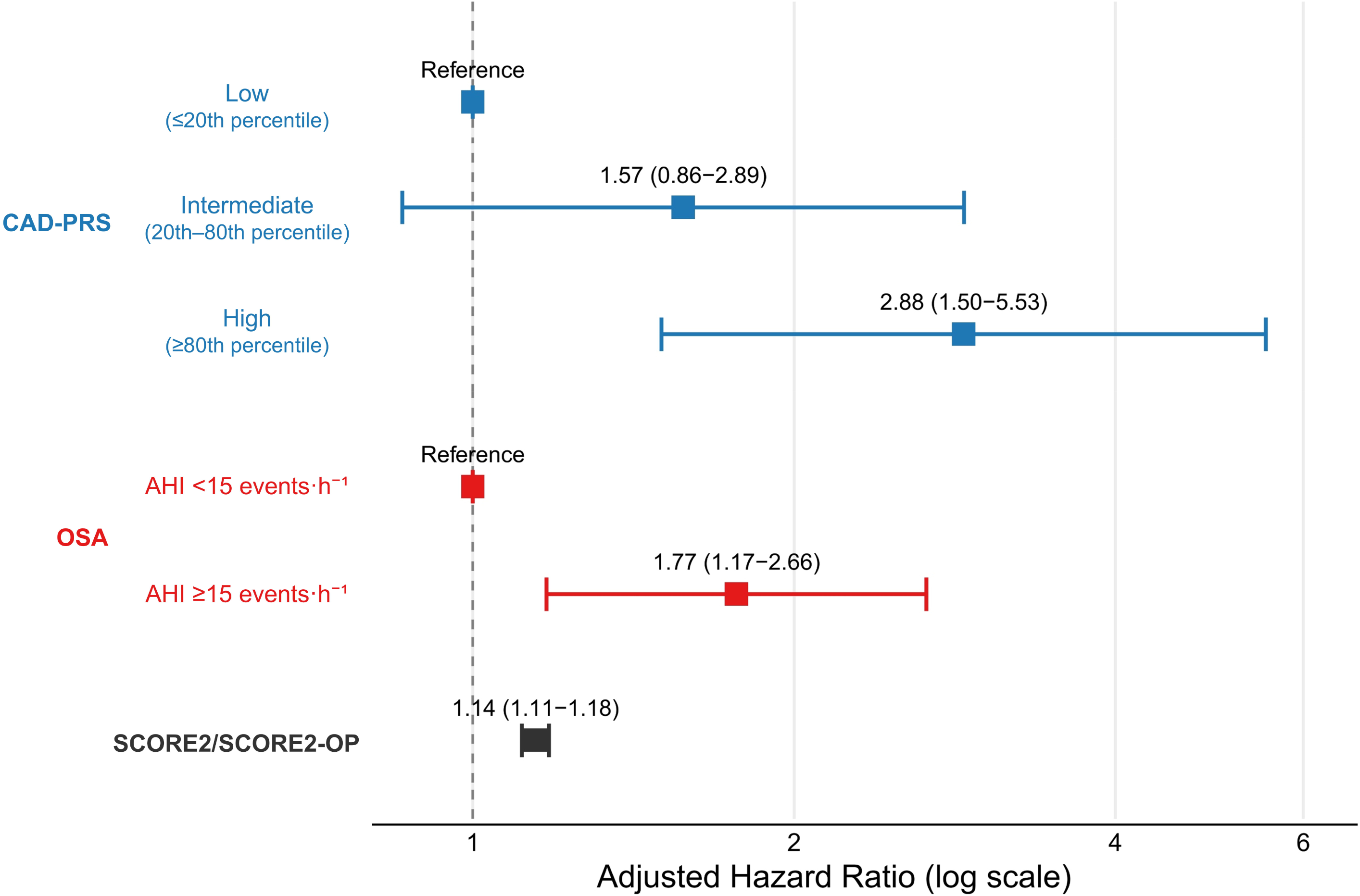
Forest plot of CAD-PRS, OSA and SCORE2 in relation to incident CV events. Forest plot showing adjusted hazard ratios and 95% confidence intervals for incident CV events from a Cox model including SCORE2/SCORE2-OP, coronary artery disease polygenic risk score (CAD-PRS) and obstructive sleep apnea (OSA). CAD-PRS is modelled in three categories: low (≤20th percentile, reference), intermediate (20th–80th percentile) and high (≥80th percentile). OSA is modelled as apnea–hypopnea index (AHI) ≥15 versus <15 events/h (reference). SCORE2/SCORE2-OP is entered as a continuous variable.

### Interaction between OSA and CAD-PRS

A significant non-linear interaction was observed between OSA severity and CAD-PRS (p-interaction = 0.013). When stratified by PRS categories, only individuals with intermediate genetic risk (PRS 20th–80th percentile) had a significantly higher CV risk associated with OSA (AHI ≥15 events/h) with a HR of 2.68 (95% CI 1.54–4.66, p-value < 0.001). In contrast, for participants at the extremes of the PRS distribution, the presence of OSA did not significantly modify CV risk. In the lowest PRS category, HR for OSA was 1.37 (95% CI 0.43-4.36, p-value = 0.59). In the highest PRS category, HR for OSA was 0.99 (95% CI 0.44-2.22, p-value = 0.98). Full model, non-linear interaction and adjusted cumulative incidence curve are shown in Figures 3a, 3b and 3c.

**Figure 3.**
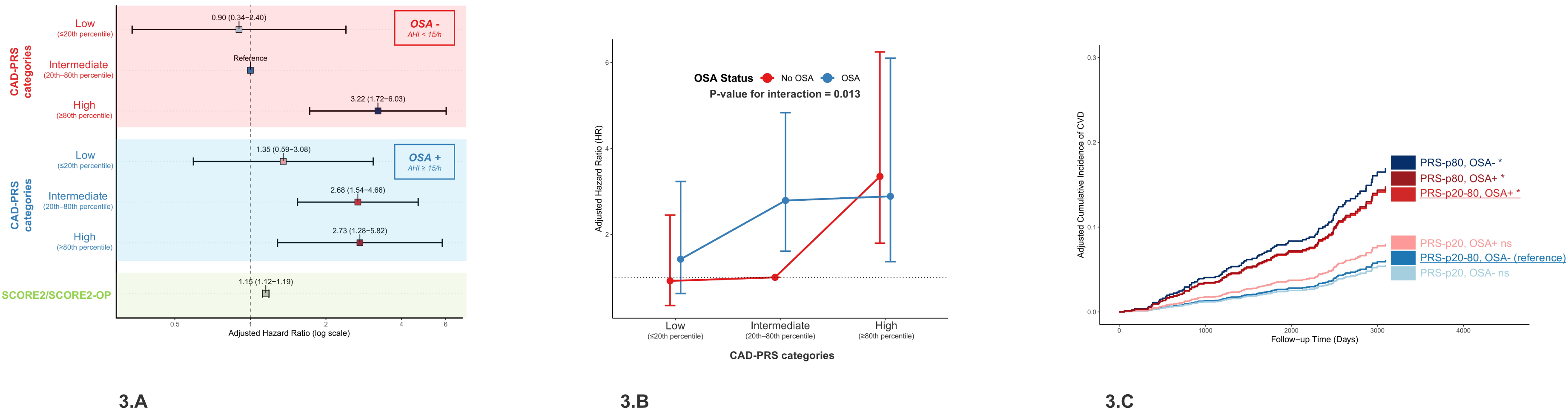
Combined effect of CAD-PRS and OSA on CV risk. (A) Forest plot showing adjusted hazard ratios (HRs) and 95% confidence intervals (CIs) for incident cardiovascular (CV) events according to coronary artery disease polygenic risk score (CAD-PRS) categories, stratified by obstructive sleep apnea (OSA) status. CAD-PRS is grouped as low (≤20th percentile), intermediate (20th–80th percentile) and high (≥80th percentile), separately for participants without OSA (apnea– hypopnea index [AHI] <15 events/h) and with moderate-to-severe OSA (AHI ≥15 events/h). (B) Interaction plot illustrating the adjusted HRs (with 95% CIs) for CV events across CAD-PRS categories in participants with and without OSA, with the corresponding p-value for the OSA×CAD-PRS interaction. (C) Adjusted cumulative incidence curves for CV events according to combined CAD-PRS/OSA groups, using the intermediate-risk, OSA-negative group as the reference. All HRs are derived from Cox model adjusted for SCORE2/SCORE2-OP

### Sensitivity analysis

Because the definition of CV outcomes may influence the observed associations, we conducted sensitivity analyses using progressively stricter outcome definitions and further adjusted the models for medication use (antidiabetic, antihypertensive, and lipid-lowering therapies). Across all outcome definitions (hard CV outcomes including TIA, hard CV outcomes excluding TIA, and pure hard CV outcomes, as described in methods) participants with moderate-to-severe OSA in the mid-range PRS category (PRS quintiles 2–4) consistently exhibited a significantly higher risk of incident CVD compared with those without OSA, consistent with the main analysis (Figures S1, S2 and S3).

The interaction between OSA and PRS remained significant for hard CV outcomes including TIA (p-value = 0.021) and excluding TIA (p-value = 0.045) and showed a similar trend for pure hard CV outcomes (p-value = 0.108) (Figure S1, S2 and S3). Further adjustment for medication use yielded essentially unchanged results, with significant interactions for any CVD (p-value = 0.016) and hard CVD (p-value = 0.048) (Figure S4 and S5). These findings highlight the robustness of the observed interaction between OSA status and genetic risk, particularly among individuals in the intermediate PRS category.

### Model performances and Improvement in risk classification

Global model performance metrics are summarized in Table 2. The base model achieved a C-index of 0.768 and a Nagelkerke’s R² of 0.088. Adding the PRS improved discrimination (C-index 0.781, R² = 0.104; LRT p-value = 0.035), while the inclusion of OSA also significantly enhanced model fit (C-index 0.78, R² = 0.112; LRT p-value = 0.004). The combined model incorporating PRS, OSA and their interaction term achieved the highest overall explanatory power (C-index 0.782, R² = 0.118; LRT p-value = 0.001), although incremental gains in discrimination were limited. All models showed good calibration (Figure S6).

To further assess the clinical relevance of these improvements, we examined reclassification metrics when compared to SCORE2 as a continuous variable (Table 3). The PRS yielded only modest gains in discrimination (IDI 0.020, p-value = 0.016) without significant improvement in net reclassification. In contrast, OSA provided substantial incremental value (IDI 0.027, p-value < 0.001; continuous NRI 0.450, p-value = 0.068). The combined model including both PRS and OSA, and particularly the model with their interaction term, achieved the strongest reclassification improvements (IDI 0.038 p-value = 0.004 and 0.057 p-value ≤ 0.001, respectively), with significant increases in continuous NRI and median risk score improvement.

When using SCORE2/SCORE2-OP categories, the extended model significantly improved patient reclassification across all age groups with a total categorical NRI: 0.1707 (95% CI: 0.0420 to 0.3047, p-value = 0.014) and a NRI for Events: 0.0300 (95% CI: -0.0893 to 0.1604), NRI for Non-Events: 0.1407 (95% CI: 0.1015 to 0.1796). With SCORE2/SCORE2-OP, 915 of the 1379 participants were classified as moderate risk, where the full model reclassified 476 of these 915 (which represents 52.02%) in the low and high-risk group (N=326 and 150 respectively).

## DISCUSSION

In this large, prospective, population-based cohort, we observed three major findings. First, moderate-to-severe OSA (AHI ≥ 15 events/h) was linked to a higher incidence of adjudicated CVD events, especially in individuals with intermediate genetic susceptibility (PRS quintiles 2–4) highlighting a clinically relevant interaction between OSA and genetic predisposition. Second, integrating OSA, CAD-PRS and their interaction into the SCORE2/SCORE2-OP algorithms significantly improved cardiovascular risk discrimination and reclassification beyond clinical risk factors alone. Finally, the CAD-PRS was strongly and independently associated with the occurrence of adjudicated CV events over a 10-year follow-up among participants with and without OSA, supporting the validity of this PRS in our study population.

### Interaction between OSA and genetic risk

The excess cardiovascular risk associated with moderate-to-severe OSA was most evident in participants at intermediate genetic risk, who represented 60% of the cohort. At the extremes of genetic risk, OSA severity had little incremental value: participants with low PRS remained at low risk regardless of OSA, while those with high PRS were already at very high risk. These findings suggest that OSA severity has additional discriminatory value particularly among individuals with intermediate genetic risk, and indicate a potential ceiling effect in risk accumulation, where the distinct contribution of OSA becomes less discernible in the presence of an extreme polygenic risk profile. The model performance and improvement in risk classification indicate that while the PRS contributes modestly on its own, OSA and its interaction with genetic risk provide the greatest incremental predictive value.

### Clinical implications

Our findings have direct clinical relevance. Incorporating CAD-PRS into existing clinical risk scores could help clinicians identify OSA patients at highest cardiovascular risk. This could support more aggressive prevention strategies, such as stricter control of modifiable risk factors, prioritization for follow-up and earlier initiation of continuous positive airway pressure (CPAP) therapy.

There is also evidence that knowledge of genetic risk can motivate positive health behaviours and treatment adherence^44,45^. Thus, CAD-PRS may not only refine risk prediction but also enhance patient engagement, which is crucial in OSA management where CPAP adherence remains a major challenge. However, future studies should investigate whether CAD-PRS can predict the cardiovascular benefit of CPAP therapy, as previously shown for statins and other preventive interventions in high genetic risk patients^46^.

Importantly, patients with moderate OSA (AHI 15–30 events/h) represent a particularly challenging group for treatment decisions, especially when they are minimally symptomatic or asymptomatic. In this population, the added value of CAD-PRS in identifying individuals at higher CV risk is encouraging, as it may help refine therapeutic decisions and supports further studies evaluating whether CPAP effectively reduces cardiovascular events in genetically high-risk moderate OSA patients.

Beyond our study, CAD-PRS are likely to become increasingly accessible as genotyping and sequencing costs fall, and large biobanks and testing platforms continue to develop. Because a PRS can be calculated once from genome-wide data and then used throughout life, it can be cost-effective even for a single indication as shown by Mujwara et al.^47^, and its use is compatible with routine care. As such, incorporating PRS into clinical decision-making may, in the future, help clinicians better stratify CV risk in patients with moderate OSA.

### Strengths and limitations

The strengths of our study include its prospective design, long follow-up, rigorous adjudication of CV events, comprehensive phenotyping with home-based PSG, and use of a validated CAD-PRS.

However, several limitations merit consideration. The number of cardiovascular events was limited, which may have reduced power for subgroup and interaction analyses. The study population consisted exclusively of individuals of European ancestry, limiting generalizability to other ethnicities. CPAP treatment and other OSA interventions during follow-up were not systematically recorded and could have influenced the observed associations. Finally, this study lacks external validation for further generalisation.

### Future directions

Our findings pave the way for further research integrating genetic information into the management of OSA. Larger, multi-ethnic cohorts are needed to validate these observations, and randomized studies should test whether CAD-PRS-guided strategies improve cardiovascular outcomes among OSA patients. In addition, evaluating whether CAD-PRS modifies the benefit of CPAP therapy or other interventions could inform more personalized treatment approaches.

## CONCLUSION

A CAD-PRS is independently associated with incident CV and interacts with OSA status to refine cardiovascular risk stratification. The excess risk conferred by moderate-to-severe OSA was particularly evident among individuals with intermediate genetic risk, who represent the majority of the population. Incorporating genetic risk and OSA status into established clinical scores such as SCORE2 significantly improved model performance and reclassification, underscoring the potential of CAD-PRS to guide cardiovascular risk assessment in OSA patients.

## Acknowledgements

The authors thank Professor Gérard Waeber, Professor Vincent Mooser, Daniela Andries, and Nadia Tobback for their important contributions to the HypnoLaus and CoLaus|PsyCoLaus cohorts, the Lausanne population that volunteered to participate in the CoLaus|PsyCoLaus and HypnoLaus studies, and the entire CoLaus|PsyCoLaus team.

## Data Availability

The data of CoLaus|PsyCoLaus used in this study cannot be fully shared as they contain potentially sensitive personal information on participants. According to the Ethics Committee, sharing these data would be a violation of the Swiss legislation concerning privacy protection. However, coded individual-level data that do not allow researchers to identify participants are available upon request to researchers who meet the criteria for data sharing (CHUV, Lausanne, Switzerland). Any researcher affiliated with a public or private research institution who complies with the CoLaus|PsyCoLaus standards can submit a research application to. Detailed instructions for gaining access to the CoLaus|PsyCoLaus data used in this study are available at www.colaus-psycolaus.ch/professionals/how-to-collaborate/.

## FIGURES TITLES AND LEGENDS

**Figure S1. Interaction between OSA and CAD-PRS for hard cardiovascular outcomes including TIA (A–C)**

Legend – Figure S1. (A) Forest plot, (B) interaction plot and (C) cumulative incidence curves for hard CV outcomes including TIA, according to OSA status and CAD-PRS. Hard CV outcomes including TIA comprise CV death, stroke (definite or probable) and TIA (excluding amaurosis fugax and transient global amnesia), AMI and CHD, definite or probable. Models are adjusted for SCORE2/SCORE2-OP; the OSA×PRS interaction is significant (p = 0.021).

Abbreviations: OSA, obstructive sleep apnea; CAD-PRS, coronary artery disease polygenic risk score; PRS, polygenic risk score; CV, cardiovascular; TIA, transient ischaemic attack; AMI, acute myocardial infarction; CHD, coronary heart disease

**Figure S2. Interaction between OSA and CAD-PRS for hard cardiovascular outcomes excluding TIA (A–C)**

Legend – Figure S2. (A) Forest plot, (B) interaction plot and (C) cumulative incidence curves for hard CV outcomes excluding TIA, according to OSA status and CAD-PRS. Hard CV outcomes excluding TIA comprise cardiovascular death, stroke (definite or probable; excluding TIA, amaurosis fugax and transient global amnesia), AMI and CHD, definite or probable. Models are adjusted for SCORE2/SCORE2-OP; the OSA×PRS interaction remains significant (p-value = 0.045).

Abbreviations: OSA, obstructive sleep apnea; CAD-PRS, coronary artery disease polygenic risk score; PRS, polygenic risk score; CV, cardiovascular; TIA, transient ischaemic attack; AMI, acute myocardial infarction; CHD, coronary heart disease

**Figure S3. Interaction between OSA and CAD-PRS for pure hard cardiovascular outcomes (A–C)**

Legend – Figure S3. (A) Forest plot, (B) interaction plot and (C) cumulative incidence curves for pure hard CV outcomes according to OSA status and CAD-PRS. Pure hard CV outcomes comprise cardiovascular death, definite stroke, definite AMI and definite CHD. Models are adjusted for SCORE2/SCORE2-OP; the OSA×PRS interaction shows a similar trend to the main analysis (p-value = 0.108).

Abbreviations: OSA, obstructive sleep apnea; CAD-PRS, coronary artery disease polygenic risk score; PRS, polygenic risk score; CV, cardiovascular; AMI, acute myocardial infarction; CHD, coronary heart disease

**Figure S4. Interaction between OSA and CAD-PRS for any cardiovascular disease after adjustment for medication (A–C)**

Legend – Figure S4. (A) Forest plot, (B) interaction plot and (C) cumulative incidence curves for any CVD according to OSA status and CAD-PRS, with additional adjustment for antidiabetic, antihypertensive and lipid-lowering therapies. Models are adjusted for SCORE2/SCORE2-OP and medication use; the OSA×PRS interaction remains significant (p-value = 0.016).

Abbreviations: OSA, obstructive sleep apnea; CAD-PRS, coronary artery disease polygenic risk score; PRS, polygenic risk score; CV, cardiovascular

**Figure S5. Interaction between OSA and CAD-PRS for hard cardiovascular disease after adjustment for medication (A–C)**

Legend – Figure S5. (A) Forest plot, (B) interaction plot and (C) cumulative incidence curves for hard CVD according to OSA status and CAD-PRS, further adjusted for antidiabetic, antihypertensive and lipid-lowering therapies. Hard CVD is defined as in Figure S2. Models are adjusted for SCORE2/SCORE2-OP and medication use; the OSA×PRS interaction remains significant (p-value = 0.048).

Abbreviations: OSA, obstructive sleep apnea; CAD-PRS, coronary artery disease polygenic risk score; PRS, polygenic risk score; CVD, cardiovascular disease; AMI, acute myocardial infarction; CHD, coronary heart disease

**Figure S6. Calibration of models for 10-year cardiovascular risk**

Legend – Figure S6. The plot illustrates the agreement between predicted 10-year risk (x-axis) and observed event frequency (y-axis). The diagonal dashed line represents perfect calibration. The gray shaded area depicts the distribution of predicted risks within the cohort, showing that the vast majority of participants fall into the low-to-intermediate risk category (<10%). All models showed good calibration, with predicted risks closely matching the ideal line of perfect agreement in the low and intermediate risk ranges, indicating no systematic bias in risk estimation. In the high-risk range (>10%), models tended to overestimate risk, reflecting the sparsity of data in the upper tail and the potential impact of preventive interventions (e.g., lipid-lowering therapy) initiated in high-risk individuals during the follow-up period.

Abbreviations: OSA, obstructive sleep apnea; PRS, polygenic risk score.

## TABLES

**Table 1. Patient Characteristics according to incident cardiovascular events**

**Table 2. Comparison of Cox models for prediction of cardiovascular events**

**Table 3. Improvements in discrimination and reclassification after adding CAD-PRS and OSA**

**Table S1. Baseline characteristics according to obstructive sleep apnea status**

## REFERENCES

1. Heinzer R, Vat S, Marques-Vidal P, et al. Prevalence of sleep-disordered breathing in the general population: the HypnoLaus study. Lancet Respir Med. 2015;3(4):310–318. doi:10.1016/S2213-2600(15)00043-0

2. Gottlieb DJ, Yenokyan G, Newman AB, et al. Prospective study of obstructive sleep apnea and incident coronary heart disease and heart failure: the sleep heart health study. Circulation. 2010;122(4):352–360. doi:10.1161/CIRCULATIONAHA.109.901801

3. Hirotsu C, Haba-Rubio J, Togeiro SM, et al. Obstructive sleep apnoea as a risk factor for incident metabolic syndrome: a joined Episono and HypnoLaus prospective cohorts study. Eur Respir J. 2018;52(5). doi:10.1183/13993003.01150-2018

4. Peppard PE, Young T, Palta M, Skatrud J. Prospective study of the association between sleep-disordered breathing and hypertension. N Engl J Med. 2000;342(19):1378–1384. doi:10.1056/NEJM200005113421901

5. Redline S, Yenokyan G, Gottlieb DJ, et al. Obstructive sleep apnea-hypopnea and incident stroke: the sleep heart health study. Am J Respir Crit Care Med. 2010;182(2):269–277. doi:10.1164/rccm.200911-1746OC

6. Pevernagie DA, Gnidovec-Strazisar B, Grote L, et al. On the rise and fall of the apnea-hypopnea index: A historical review and critical appraisal. J Sleep Res. 2020;29(4):e13066. doi:10.1111/jsr.13066

7. McEvoy RD, Antic NA, Heeley E, et al. CPAP for Prevention of Cardiovascular Events in Obstructive Sleep Apnea. N Engl J Med. 2016;375(10):919–931. doi:10.1056/NEJMoa1606599

8. Peker Y, Glantz H, Eulenburg C, Wegscheider K, Herlitz J, Thunström E. Effect of Positive Airway Pressure on Cardiovascular Outcomes in Coronary Artery Disease Patients with Nonsleepy Obstructive Sleep Apnea. The RICCADSA Randomized Controlled Trial. Am J Respir Crit Care Med. 2016;194(5):613–620. doi:10.1164/rccm.201601-0088OC

9. Sánchez-de-la-Torre M, Sánchez-de-la-Torre A, Bertran S, et al. Effect of obstructive sleep apnoea and its treatment with continuous positive airway pressure on the prevalence of cardiovascular events in patients with acute coronary syndrome (ISAACC study): a randomised controlled trial. Lancet Respir Med. 2020;8(4):359–367. doi:10.1016/S2213-2600(19)30271-1

10. Heinzer R, Eckert D. Treatment for obstructive sleep apnoea and cardiovascular diseases: are we aiming at the wrong target? Lancet Respir Med. 2020;8(4):323–325. doi:10.1016/S2213-2600(19)30351-0

11. Malhotra A, Ayappa I, Ayas N, et al. Metrics of sleep apnea severity: beyond the apnea-hypopnea index. Sleep. 2021;44(7). doi:10.1093/sleep/zsab030

12. Azarbarzin A, Sands SA, Stone KL, et al. The hypoxic burden of sleep apnoea predicts cardiovascular disease-related mortality: the Osteoporotic Fractures in Men Study and the Sleep Heart Health Study. Eur Heart J. 2019;40(14):1149–1157. doi:10.1093/eurheartj/ehy624

13. Butler MP, Emch JT, Rueschman M, et al. Apnea-Hypopnea Event Duration Predicts Mortality in Men and Women in the Sleep Heart Health Study. Am J Respir Crit Care Med. 2019;199(7):903–912. doi:10.1164/rccm.201804-0758OC

14. Azarbarzin A, Sands SA, Younes M, et al. The Sleep Apnea-Specific Pulse-Rate Response Predicts Cardiovascular Morbidity and Mortality. Am J Respir Crit Care Med. 2021;203(12):1546–1555. doi:10.1164/rccm.202010-3900OC

15. Blanchard M, Imler T, Hu WH, et al. Heart rate response and cardiovascular risk during obstructive sleep apnoea: an easy biomarker derived from pulse oximetry. Eur Respir J. 2025;65(5):2401883. doi:10.1183/13993003.01883-2024

16. Solelhac G, Sánchez-de-la-Torre M, Blanchard M, et al. Pulse Wave Amplitude Drops Index: A Biomarker of Cardiovascular Risk in Obstructive Sleep Apnea. Am J Respir Crit Care Med. 2023;207(12):1620–1632. doi:10.1164/rccm.202206-1223OC

17. Mazzotti DR, Keenan BT, Lim DC, Gottlieb DJ, Kim J, Pack AI. Symptom Subtypes of Obstructive Sleep Apnea Predict Incidence of Cardiovascular Outcomes. Am J Respir Crit Care Med. 2019;200(4):493–506. doi:10.1164/rccm.201808-1509OC

18. Lechat B, Appleton S, Melaku YA, et al. The association of co-morbid insomnia and sleep apnea with prevalent cardiovascular disease and incident cardiovascular events. J Sleep Res. 2022;31(5):e13563. doi:10.1111/jsr.13563

19. Solelhac G, Wachinou AP, Goyal A, et al. Prevalence and clinical significance of comorbid insomnia and sleep apnea (COMISA) in three population-based cohorts from Benin, Switzerland and India. Sleep Med. 2025;131:106526. doi:10.1016/j.sleep.2025.106526

20. SCORE2 working group and ESC Cardiovascular risk collaboration. SCORE2 risk prediction algorithms: new models to estimate 10-year risk of cardiovascular disease in Europe. Eur Heart J. 2021;42(25):2439–2454. doi:10.1093/eurheartj/ehab309

21. SCORE2-OP working group and ESC Cardiovascular risk collaboration. SCORE2-OP risk prediction algorithms: estimating incident cardiovascular event risk in older persons in four geographical risk regions. Eur Heart J. 2021;42(25):2455–2467. doi:10.1093/eurheartj/ehab312

22. Kurniansyah N, Strausz SJ, Chittoor G, et al. Polygenic scores for obstructive sleep apnoea reveal pathways contributing to cardiovascular disease. eBioMedicine. 2025;117:105790. doi:10.1016/j.ebiom.2025.105790

23. Inouye M, Abraham G, Nelson CP, et al. Genomic Risk Prediction of Coronary Artery Disease in 480,000 Adults. JACC. 2018;72(16):1883–1893. doi:10.1016/j.jacc.2018.07.079

24. Goodman MO, Cade BE, Shah N, et al. Pathway-specific Polygenic Risk Scores (PRS) Identify OSA-related Pathways Differentially Moderating Genetic Susceptibility to CAD. Circ Genomic Precis Med. 2022;15(5):e003535. doi:10.1161/CIRCGEN.121.003535

25. Firmann M, Mayor V, Vidal PM, et al. The CoLaus study: a population-based study to investigate the epidemiology and genetic determinants of cardiovascular risk factors and metabolic syndrome. BMC Cardiovasc Disord. 2008;8:6. doi:1471-2261-8-6%20%5Bpii%5D%2010.1186/1471-2261-8-6

26. Beuret H, Hausler N, Nanchen D, Mean M, Marques-Vidal P, Vaucher J. Comparison of Swiss and European risk algorithms for cardiovascular prevention in Switzerland. Eur J Prev Cardiol. 2021;28(2):204–210. doi:10.1177/2047487320906305

27. Buysse DJ, Reynolds CF, Monk TH, Berman SR, Kupfer DJ. The Pittsburgh Sleep Quality Index: a new instrument for psychiatric practice and research. Psychiatry Res. 1989;28(2):193–213.

28. Johns MW. A new method for measuring daytime sleepiness: the Epworth sleepiness scale. Sleep. 1991;14(6):540–545.

29. Iber C AIS. The AASM Manual for the scoring of sleep and associated events : Rules, terminology and technical specifications. 1st Ed Westchest Ill Am Acad Sleep Med. 2007.

30. Berry RB, Budhiraja R, Gottlieb DJ, et al. Rules for scoring respiratory events in sleep: update of the 2007 AASM Manual for the Scoring of Sleep and Associated Events. Deliberations of the Sleep Apnea Definitions Task Force of the American Academy of Sleep Medicine. J Clin Sleep Med JCSM Off Publ Am Acad Sleep Med. 2012;8(5):597–619. doi:10.5664/jcsm.2172

31. McCarthy S, Das S, Kretzschmar W, et al. A reference panel of 64,976 haplotypes for genotype imputation. Nat Genet. 2016;48(10):1279–1283. doi:10.1038/ng.3643

32. Durbin R. Efficient haplotype matching and storage using the positional Burrows-Wheeler transform (PBWT). Bioinforma Oxf Engl. 2014;30(9):1266–1272. doi:10.1093/bioinformatics/btu014

33. Loh PR, Danecek P, Palamara PF, et al. Reference-based phasing using the Haplotype Reference Consortium panel. Nat Genet. 2016;48(11):1443–1448. doi:10.1038/ng.3679

34. Manichaikul A, Mychaleckyj JC, Rich SS, Daly K, Sale M, Chen WM. Robust relationship inference in genome-wide association studies. Bioinforma Oxf Engl. 2010;26(22):2867–2873. doi:10.1093/bioinformatics/btq559

35. Chang CC, Chow CC, Tellier LC, Vattikuti S, Purcell SM, Lee JJ. Second-generation PLINK: rising to the challenge of larger and richer datasets. GigaScience. 2015;4(1):s13742-015-0047-0048. doi:10.1186/s13742-015-0047-8

36. Choi SW, O’Reilly PF. PRSice-2: Polygenic Risk Score software for biobank-scale data. GigaScience. 2019;8(7):giz082. doi:10.1093/gigascience/giz082

37. Lambert SA, Gil L, Jupp S, et al. The Polygenic Score Catalog as an open database for reproducibility and systematic evaluation. Nat Genet. 2021;53(4):420–425. doi:10.1038/s41588-021-00783-5

38. Perez-Vicencio D, Doudesis D, Thurston AJ, Selva J. RiskScorescvd: Cardiovascular Risk Scores Calculator. June 2023:0.3.1. doi:10.32614/CRAN.package.RiskScorescvd

39. Mega JL, Stitziel NO, Smith JG, et al. Genetic risk, coronary heart disease events, and the clinical benefit of statin therapy: an analysis of primary and secondary prevention trials. The Lancet. 2015;385(9984):2264–2271. doi:10.1016/S0140-6736(14)61730-X

40. Marston NA, Pirruccello JP, Melloni GEM, et al. Predictive Utility of a Coronary Artery Disease Polygenic Risk Score in Primary Prevention. JAMA Cardiol. 2023;8(2):130–137. doi:10.1001/jamacardio.2022.4466

41. Jr FEH. rms: Regression Modeling Strategies. October 2025. https://cran.r-project.org/web/packages/rms/. Accessed November 21, 2025.

42. Ozenne B, Sørensen AL, Scheike T, Torp-Pedersen C, Gerds TA. riskRegression: Predicting the Risk of an Event using Cox Regression Models. R J. 2017;9(2):440–460. doi:10.32614/RJ-2017-062

43. Uno H, Cai T. survIDINRI: IDI and NRI for Comparing Competing Risk Prediction Models with Censored Survival Data. April 2022. https://cran.r-project.org/web/packages/survIDINRI/index.html. Accessed November 21, 2025.

44. Viigimaa M, Jürisson M, Pisarev H, et al. Effectiveness and feasibility of cardiovascular disease personalized prevention on high polygenic risk score subjects: a randomized controlled pilot study. Eur Heart J Open. 2022;2(6):oeac079. doi:10.1093/ehjopen/oeac079

45. Widén E, Junna N, Ruotsalainen S, et al. How Communicating Polygenic and Clinical Risk for Atherosclerotic Cardiovascular Disease Impacts Health Behavior: an Observational Follow-up Study. Circ Genomic Precis Med. 2022;15(2):e003459. doi:10.1161/CIRCGEN.121.003459

46. Natarajan P, Young R, Stitziel NO, et al. Polygenic Risk Score Identifies Subgroup With Higher Burden of Atherosclerosis and Greater Relative Benefit From Statin Therapy in the Primary Prevention Setting. Circulation. 2017;135(22):2091–2101. doi:10.1161/CIRCULATIONAHA.116.024436

47. Mujwara D, Henno G, Vernon ST, et al. Integrating a Polygenic Risk Score for Coronary Artery Disease as a Risk-Enhancing Factor in the Pooled Cohort Equation: A Cost-Effectiveness Analysis Study. J Am Heart Assoc. June 2022. doi:10.1161/JAHA.121.025236

