## Supplementary figures and images for "A polygenic risk score improves the prediction of cardiovascular risk associated with obstructive sleep-apnea"

### Supplementary Figures

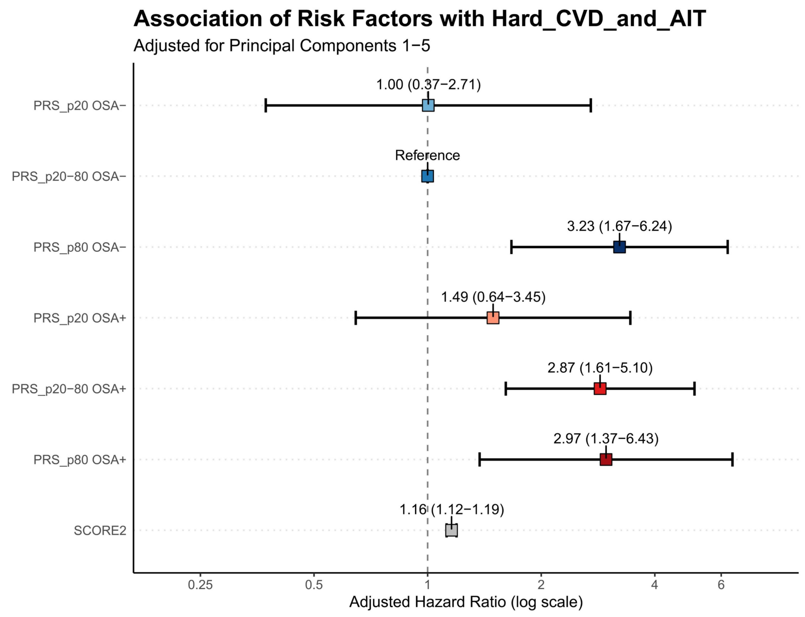

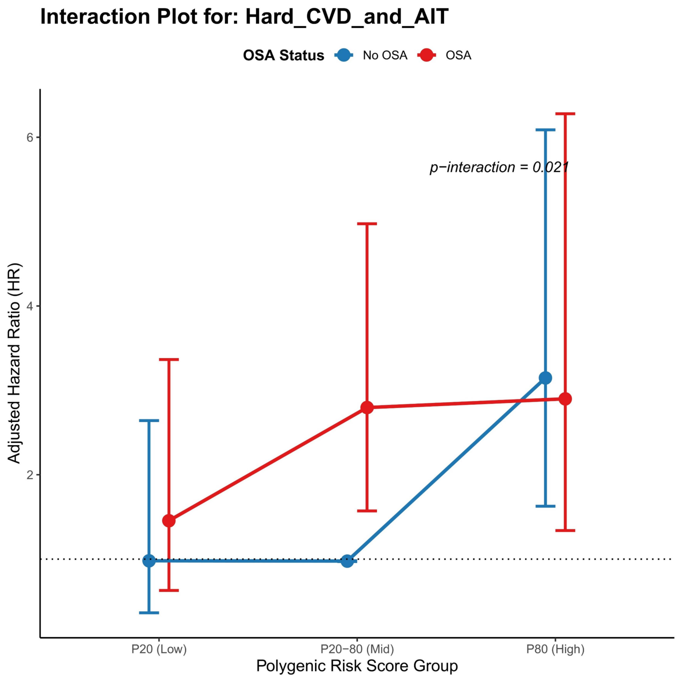

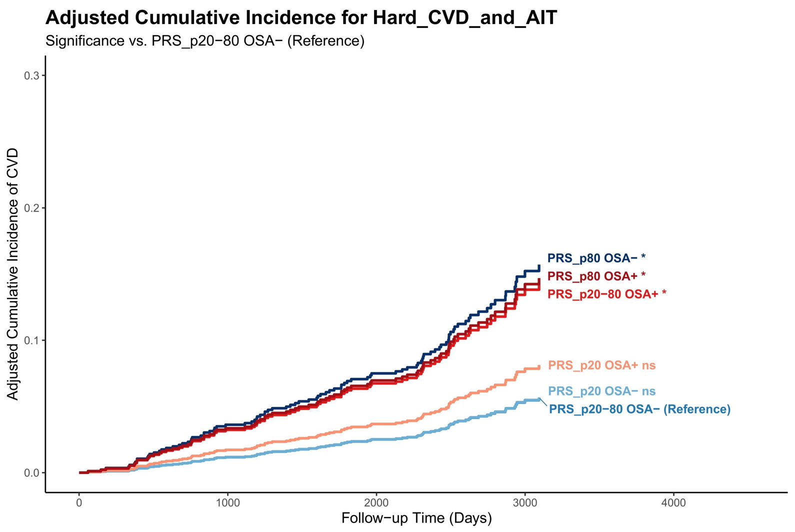

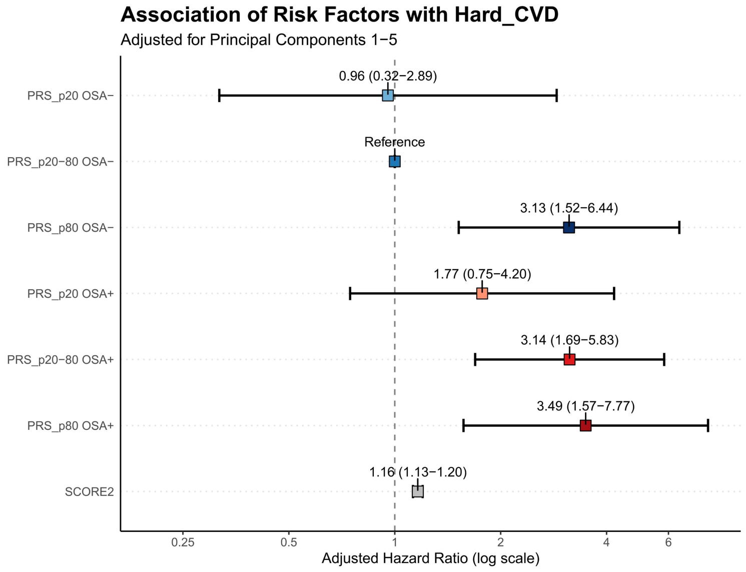

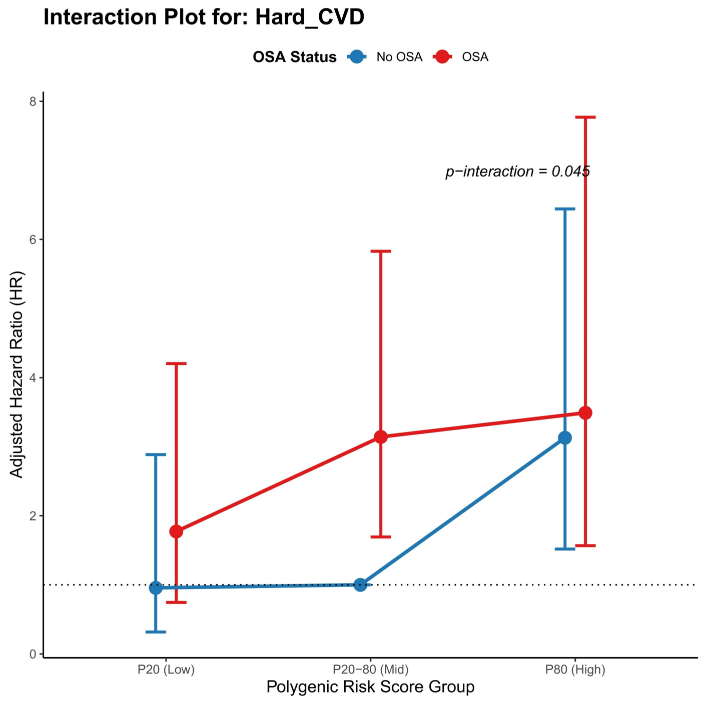

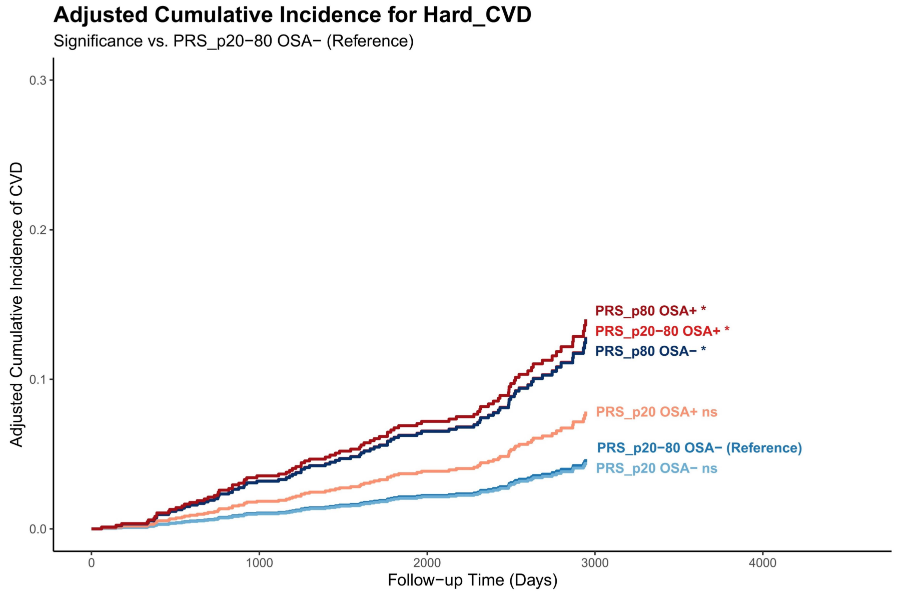

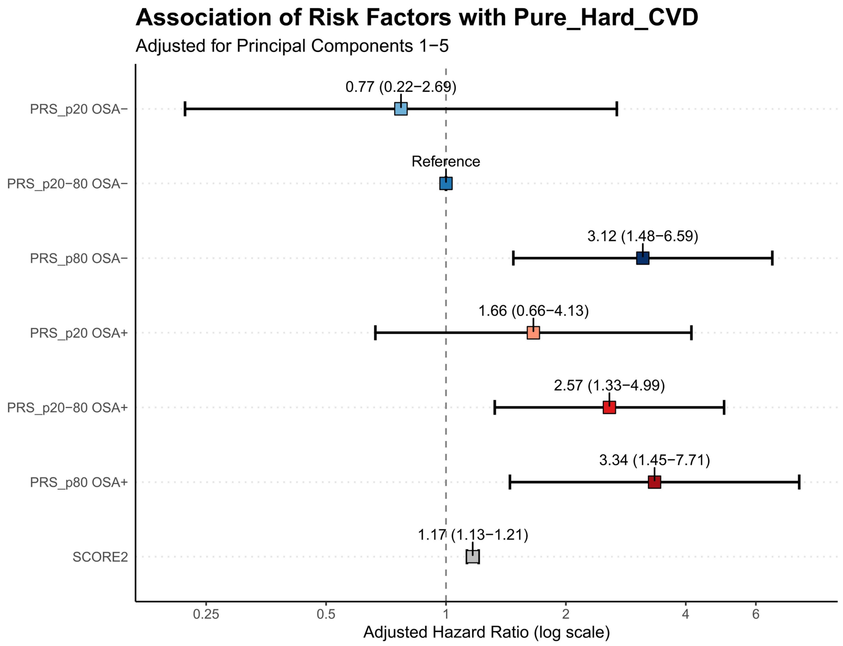

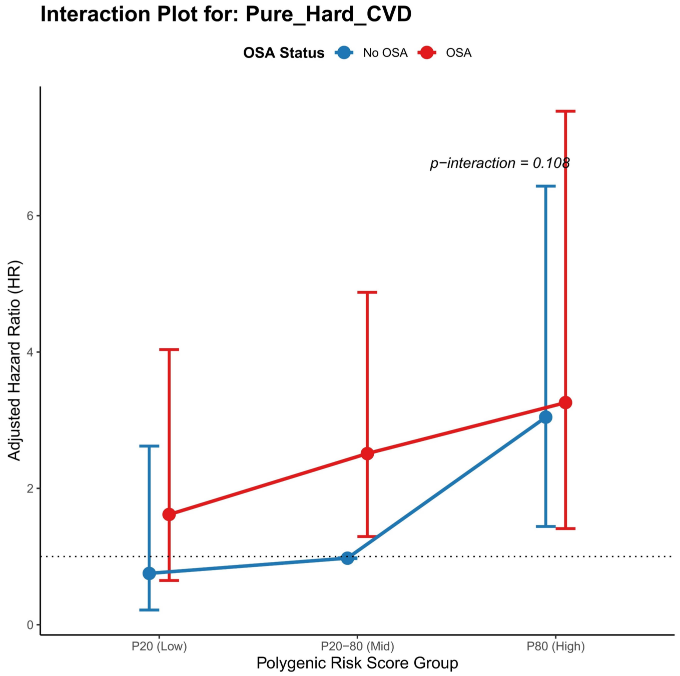

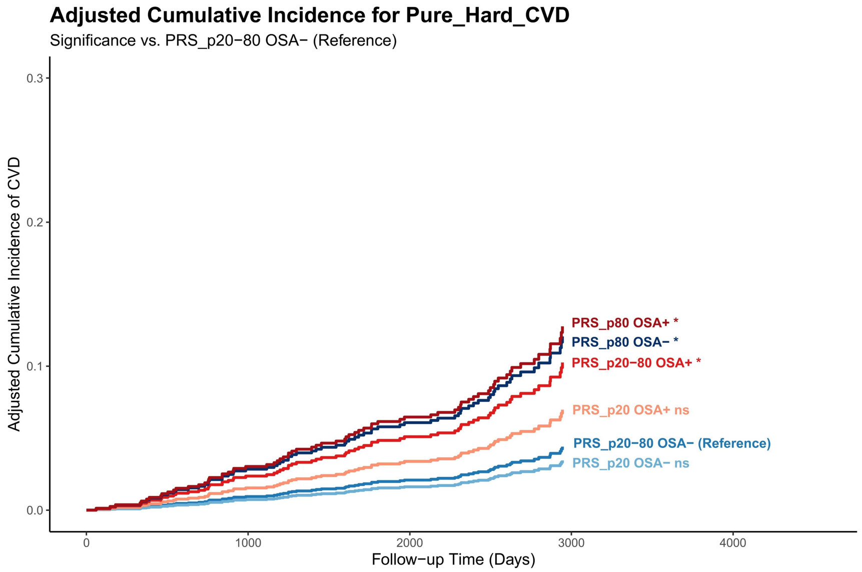

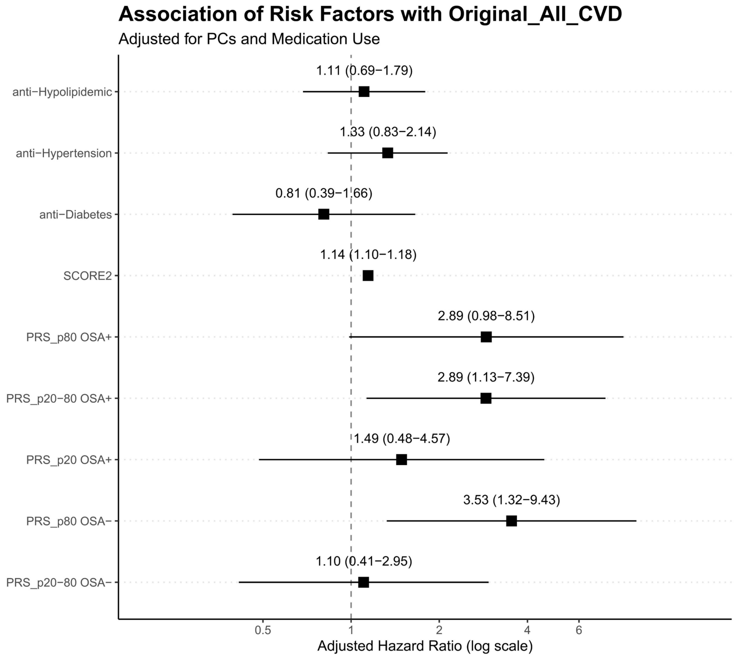

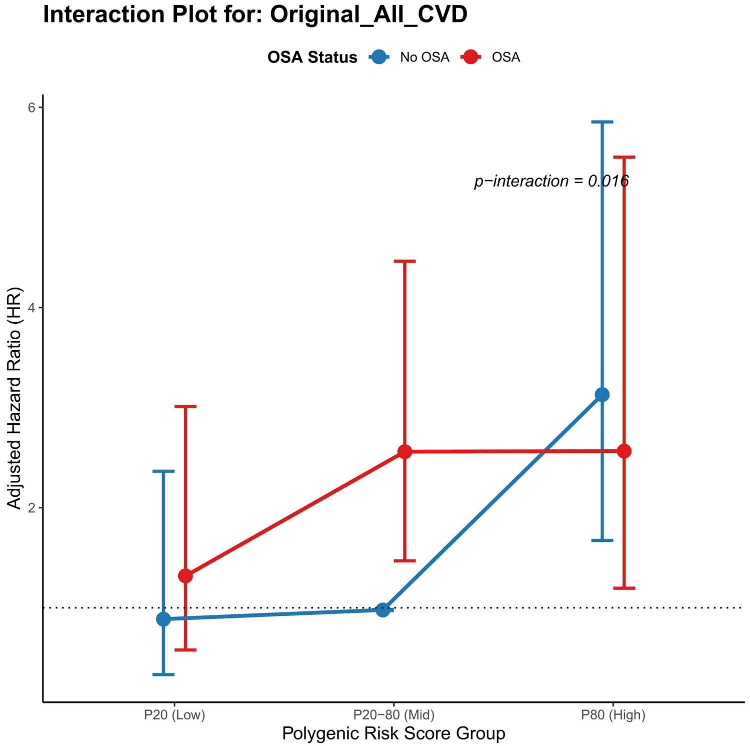

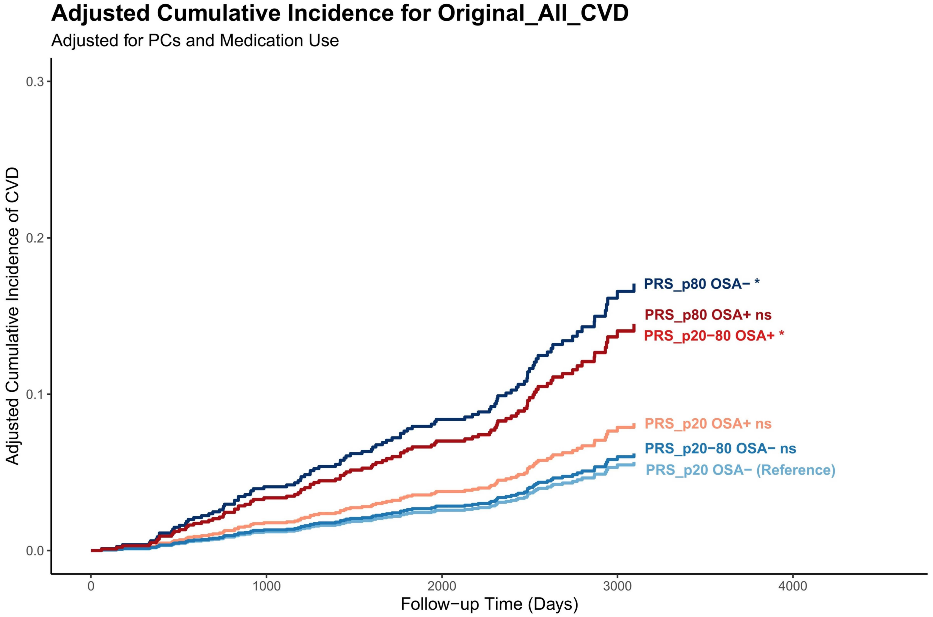

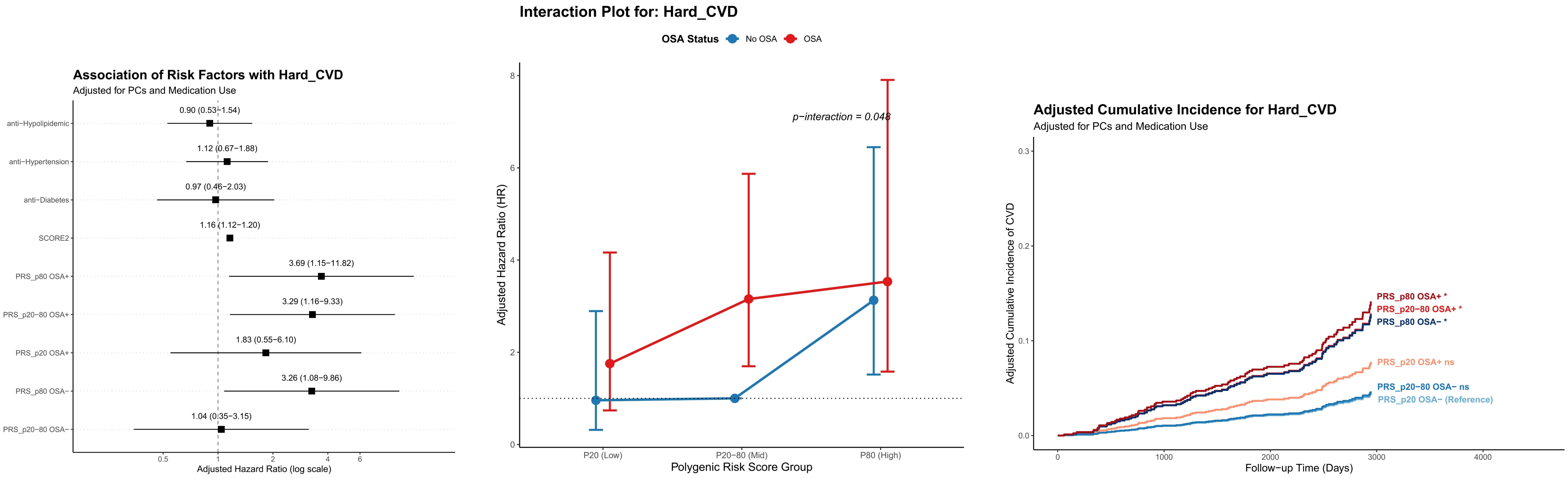

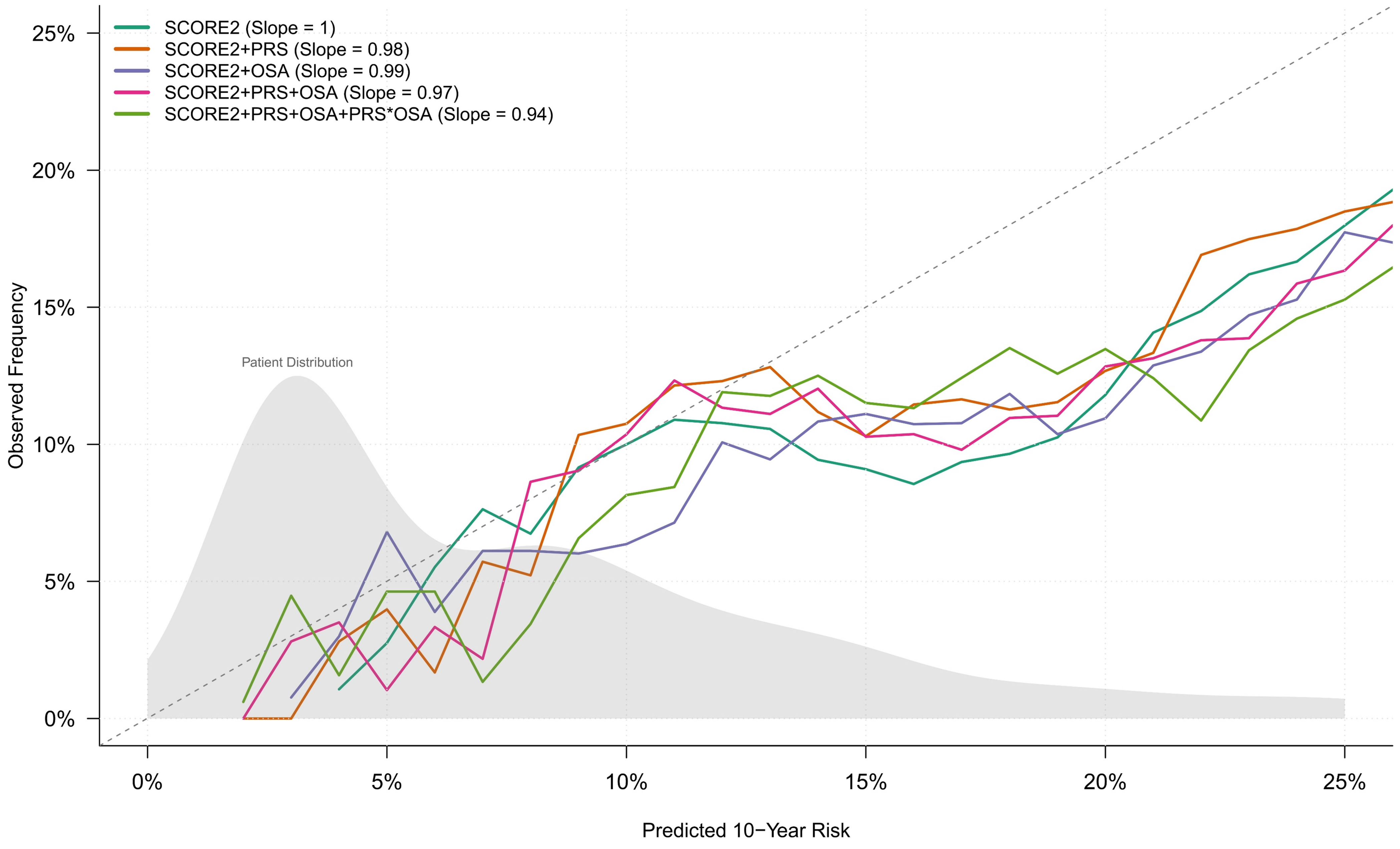
