## Supplementary Tables for "A polygenic risk score improves the prediction of cardiovascular risk associated with obstructive sleep-apnea"

### Table S1. Baseline characteristics according to obstructive sleep apnea status

| | | | OSA Status | | |
|---|---|---|---|---|---|
| **Variable** | **N** | **Overall N = 1,379*** | **AHI <15 events·h$^{-1}$ N = 891*** | **AHI ≥15 events·h$^{-1}$ N = 488*** | **p-value**** |
| Incident CV event | 1,379 | 100 (7.3%) | 45 (5.1%) | 55 (11%) | <0.001 |
| SCORE2 category | 1,379 | | | | <0.001 |
| Low risk | | 768 (56%) | 592 (66%) | 176 (36%) | |
| Moderate risk | | 472 (34%) | 245 (27%) | 227 (47%) | |
| High risk | | 139 (10%) | 54 (6.1%) | 85 (17%) | |
| Male | 1,379 | | | | <0.001 |
| No | | 731 (53%) | 561 (63%) | 170 (35%) | |
| Yes | | 648 (47%) | 330 (37%) | 318 (65%) | |
| Age (y) | 1,379 | 58 (50, 69) | 55 (48, 67) | 65 (55, 71) | <0.001 |
| BMI (kg/m2) | 1,373 | 25.6 (23.0, 28.3) | 24.6 (22.4, 27.1) | 27.3 (25.2, 30.2) | <0.001 |
| Unknown | | 6 | 2 | 4 | |
| Current smoker | 1,379 | | | | 0.24 |
| No | | 1,133 (82%) | 724 (81%) | 409 (84%) | |
| Yes | | 246 (18%) | 167 (19%) | 79 (16%) | |
| Diabetes | 1,379 | | | | <0.001 |
| No | | 1,261 (91%) | 847 (95%) | 414 (85%) | |
| Yes | | 118 (8.6%) | 44 (4.9%) | 74 (15%) | |
| Systolic BP (mmHg) | 1,379 | 125 (114, 138) | 122 (111, 135) | 130 (121, 142) | <0.001 |
| Diastolic BP (mmHg) | 1,379 | 78 (71, 85) | 77 (70, 84) | 80 (74, 88) | <0.001 |
| Hypertension | 1,379 | | | | <0.001 |
| No | | 823 (60%) | 607 (68%) | 216 (44%) | |
| Yes | | 556 (40%) | 284 (32%) | 272 (56%) | |
| Dyslipidemia | 1,379 | | | | <0.001 |
| No | | 1,030 (75%) | 695 (78%) | 335 (69%) | |
| Yes | | 349 (25%) | 196 (22%) | 153 (31%) | |
| Statins | 1,379 | | | | <0.001 |
| No | | 1,175 (85%) | 791 (89%) | 384 (79%) | |
| Yes | | 204 (15%) | 100 (11%) | 104 (21%) | |
| HDL Cholesterol (mmol/L) | 1,379 | 1.60 (1.30, 1.90) | 1.70 (1.40, 2.00) | 1.50 (1.20, 1.80) | <0.001 |
| Total Cholesterol (mmol/L) | 1,379 | 5.70 (5.00, 6.40) | 5.70 (5.00, 6.40) | 5.80 (5.10, 6.40) | 0.071 |
| Weekly alcohol consumption (units) | 1,379 | 4 (1, 10) | 4 (1, 8) | 6 (2, 12) | <0.001 |
| AHI (events/h) | 1,379 | 10 (4, 21) | 6 (3, 9) | 27 (20, 40) | <0.001 |
| ODI-3% | 1,379 | 10 (4, 19) | 6 (3, 10) | 25 (18, 37) | <0.001 |
| Total sleep time (min) | 1,379 | 405 (359, 449) | 408 (365, 454) | 399 (350, 441) | 0.003 |
| Sleep efficiency (%) | 1,379 | 88 (80, 93) | 90 (82, 93) | 84 (75, 91) | <0.001 |
| Mean SpO2 | 1,379 | 94.20 (93.10, 95.30) | 94.80 (93.80, 95.60) | 93.40 (92.25, 94.30) | <0.001 |
| PWADi | 1,369 | 52 (38, 66) | 52 (38, 65) | 52 (39, 66) | 0.23 |
| Unknown | | 10 | 4 | 6 | |

**Table S1 caption:** Baseline characteristics of the 1,379 participants according to obstructive sleep apnea (OSA) status (AHI <15 vs ≥15 events/h)

**Abbreviations:** OSA: obstructive sleep apnea; CV: cardiovascular; SCORE2: Systematic COronary Risk Evaluation 2; BMI: body mass index; BP: blood pressure; HDL: high-density lipoprotein; AHI: apnea–hypopnea index; ODI-3%: oxygen desaturation index (3% desaturation threshold); $SpO_2$: peripheral oxygen saturation by pulse oximetry; PWADi: pulse wave amplitude drops index.

***** Median (IQR) or frequency (%).

****** P-values derived from Pearson's chi-squared test for categorical variables or Wilcoxon rank-sum test for continuous variables.
